# A systematic review protocol for estimation of the prevalence of depression using diagnostic instruments in the elderly population in India, 2000-2019

**DOI:** 10.1101/2020.04.10.20060871

**Authors:** Priyamadhaba Behera, Manju Pilania, Vikas Yadav, Mohan Bairwa, Deepti Dabar, Surama Manjari Behera, S. Poongothai, Viswanathan Mohan, Shiv Dutt Gupta

## Abstract

**Introduction:** Depression is a common mental disorder in the elderly population, which significantly impacts their quality of life. However, correct estimates of its magnitude are not available in the elderly in India. The present systematic review and meta-analysis would attempt to estimate the prevalence of depression using diagnostic instruments among elderly persons aged 60 years and above.

**Methods and analysis:** Searches will be performed in PubMed, Scopus, Embase, Web of Science, CINAHL, and PsycINFO. Community-based cross-sectional and cohort studies (2001 – 9/2019) reporting the prevalence of depression in the elderly; using diagnostic instruments will be included. Studies conducted among chronic disease patients, in-hospital patients, and special groups such as with disaster-stricken populations, and studies reporting the only 1 or 2 subcategories of depression, will be excluded. Disagreements in study selection and data abstraction will be resolved by consensus and arbitration by a third reviewer. AXIS critical appraisal tool will be used for quality assessment of individual studies. Findings of eligible studies will be pooled using fixed-effects or random-effects meta-analysis whichever is appropriate. Heterogeneity between studies will be examined by Cochran’s Q test and quantified by I^2^ statistic. A cumulative meta-analysis will be used to detect temporal trends in the prevalence of depression and the effect of poor-quality studies on the pooled estimate. Publication bias will be assessed by visual inspection of funnel plots and the Egger test.

**Ethics and dissemination:** No ethical approval will be needed because it will be a systematic review. Data from previously published studies will be retrieved and analyzed. Findings will be disseminated through a peer-reviewed publication in a scientific journal and conferences.

**PROSPERO:** registration number CRD42019138453.

**Strengths and limitations of this study:**

- It is a first-ever systematic review of depression prevalence in India based on diagnostic instruments only.
- Meta-analytic techniques such as cumulative meta-analysis, leave-one-out (jack-knife estimation) meta-analysis, and meta-regression will enrich the analysis and provide the estimate of prevalence nearer to the population estimate.
- A comprehensive synthesis of all available depression prevalence data in India using a standardized risk of bias tool.
- The protocol adheres to Preferred Reporting Items for Systematic Reviews and Meta-Analyses Protocols guidelines.
- Heterogeneity in methodologies, such as diagnostic criteria, study duration, sampling design, and study locations may limit comparison across studies.

## Introduction

Mental disorders have emerged as one of the major health problems in India and globally. Being chronic in nature, they significantly impact the quality of life (1–3). Depression is the most common mental disorder affecting 322 million people globally, with a prevalence ranging from 4 to 13% for minor depression and 1 to 4% for major depression (4–6). Depression affects not only the quality of life but also increases the risk of all-cause mortality, including cardiovascular diseases and stroke (7). The Global Burden of Disease study projected that depression will be the leading cause of Disability-Adjusted Life Years (DALY) in developing countries by 2020 (8). Depression has emerged out as a significant risk factor for suicidality and suicide deaths in India (9). With the rapid population aging, the proportion of elderly persons is estimated to increase from 12% to 22% between 2015 and 2050 in the world. In India, the figure will rise from the current 8.6% in 2011 to 19% by 2050 (10,11). This underscores the significant health burden depression will place on elderly people in India in the years to come. In India, “elderly persons” are those who have attained the age of 60 years and above (12,13).

Depression is both an underdiagnosed and undertreated mental disorder in elderly persons, and its varied presentation makes its diagnosis difficult. Elderly persons with depression have poorer functioning as compared to people in a similar age group without depression and have increased health care costs (6,14). Even though India’s population is rapidly aging, little is known about the magnitude of depression at the national and regional levels. The estimated prevalence of depression among elderly persons from rural community-based studies of India varied highly from 12.7% to 53.7% (15). With this background, we attempted to estimate the prevalence of depression in the elderly population in India, using published studies that employed standardized screening tests to identify depression (16). That study could have provided a higher estimate of the depression prevalence in elderly people as the screening tests might have overestimated the prevalence, given the higher sensitivity of the screening tests, albeit their low specificity. Indeed, the screening tests blur the distinctions between low and high prevalence populations due to false positives (17–19).

Moreover, the prevalence studies vary in methodologies, including variable sensitivity and specificity of screening of tests, geographical and cultural characteristics, and level of expertise among the investigators (20–22). Nevertheless, these studies indicated that the prevalence of depression should be estimated using reliable and validated diagnostic tools to identify depression more accurately and to help with planning and health systems management (16,19). A comprehensive clinical interview using a sensitive and specific diagnostic tool is the gold standard for confirming a diagnosis of depression, which also helps plan the appropriate therapy (23). Therefore, we designed this systematic review and meta-analysis to estimate the prevalence of depression by including the studies that have used diagnostic instruments among elderly persons in India.

## Methods

The protocol has been prepared following the Preferred Reporting Items for Systematic Reviews and Meta-analysis Protocol (PRISMA-P) (24), which provides a standardized guide for performing systematic reviews and meta-analysis (Appendix 1. PRISMA-P checklist). The protocol has been registered on PROSPERO (CRD42019138453) (25).

### Eligibility criteria

The elderly population aged 60 years and above in India is the population of interest. This review will include the studies with the following eligibility criteria:

#### Inclusion criteria

1. Community-based cross-sectional and cohort studies published during 01/2001 – 9/2019
2. Studies that reported the prevalence of depression/ depressive symptoms using diagnostic instruments for identifying depression. “Diagnostic instruments” are tools that diagnose depression by the International Classification of Diseases criteria and/or Diagnostic and Statistical Manual of Mental Disorders criteria (26).

#### Exclusion criteria

1. Studies among elderly patients with chronic diseases such as diabetes, HIV/AIDS, etc, in-hospital patients, and other special groups such as with disaster-stricken experiences.
2. Studies which reported either 1 or 2 subcategories of depression (mild, moderate and severe) only
3. Studies in which unstructured clinician-defined diagnosis

### Information sources

Searches will be performed in PubMed, Scopus, Embase, Web of Science, CINAHL, and PsycINFO. To enrich and supplement the literature search, the references of selected articles and relevant reviews will be scanned. Then, we will circulate a list of identified articles to the systematic review team, as well as to the selected experts working in this field to ensure the completeness of search results.

### Search strategy

Initially, controlled descriptors (such as MeSH terms, CINAHL headings, PsycINFO thesaurus) will be identified in each database. Following keywords such as “psychiatric”, “depression”, “mental”, “depressive disorders”, “aged”, “geriatric”, “elderly”, “old aged”, “aging”, “prevalence”, “epidemiological studies”, “epidemiology”, and “India” will be used to develop the search strategy. Appropriate Boolean operators will be employed. We will not impose any language filter. The search will be limited to human subjects. The search strategy for PubMed is given below:

**#1**. psychiatric OR depressi* OR mental OR “Depression”[Mesh] OR “Depressive Disorder”[Mesh]

**#2**. “Aged”[Mesh] OR geriatric OR elder* OR “old aged” OR aging

**#3**. “Prevalence”[Mesh] OR prevalence OR “Epidemiology”[Mesh] OR “epidemiological stud*”

**#4**. India

**#5**. #1 AND #2 AND #3 AND #4

**#6**. Filters: Publication date from 2001/01/01, Humans.

### Selection process and data management

Two reviewers (MP and PMB) will conduct searches in all identified databases. All search results will be imported into Rayyan QCRI Software to ensure a systematic and comprehensive search and document the selection process (27). Another reviewer (VY) will manage the Rayyan and identify and remove the duplicate citations and ensure an independent review of titles and abstracts by blinding the decisions of both reviewers. MP and PMB will review of titles and abstracts of the shortlisted citations in the Rayyan using a customized inclusion/exclusion checklist (population-based studies; depression prevalence, study duration, and India). After that, VY will identify the discrepancies between the two reviewers in the Rayyan software and inform them of making a consensus for the selection of the study. Full-text copies of all studies selected will be obtained to find more details. Both reviewers will review the full-text copies of articles to identify whether diagnostic instruments have been used to identify depression in the study participants.

We will record the reasons for the exclusion of all the studies for which we had obtained full copies. Wherever the studies have been reported in multiple publications/reports, all papers will be obtained. While the studies will be included as only one in the review, the data will be extracted from all the publications to ensure the maximal relevant data is retrieved. The full-text copies of all selected articles will be evaluated for quality assessment and data extraction. The study selection process will be presented using the PRISMA flow chart describing the reasons for the exclusion for the studies we will explore full texts.

The reference management software Mendeley Desktop for Windows will be used to store, organize, cite, and manage all the selected references (28).

### Data extraction

PMB and MP will independently perform data extraction on the key variables including study details (author, year of publication); methods (study design, study location, study setting, sample size, sampling method, non-response, age, sex, screening procedure, screening for dementia, diagnostic instrument); and results (risk factors of depression studied and prevalence data) will be extracted. Any disagreement in the data abstraction will be resolved by consensus, and if required, the arbitration will be done by the members of the review team (MB, VM, and SDG). First or corresponding authors will be contacted if additional information is required in the selected articles.

### Risk of bias in individual studies

AXIS critical appraisal tool will be used for quality assessment of the individual studies (29). The AXIS tool would emphasize mainly on the presented methods and results. The AXIS tool contains a 20-point questionnaire with “yes”, “no”, and “don’t know” answer that addresses study quality and reporting. The critical areas in the AXIS tool included are study design, sample size justification, target population, sampling frame, sample selection, measurement validity and reliability, overall methods, and conflict of interest and ethical issues.

### Strategy for data synthesis

In this systematic review, extracted data will be presented in comprehensive tables and flowcharts. The pooling of prevalence will be done using meta-analysis. In case the relevant information is not available for meta-analysis, a narrative synthesis will be performed. The effect size of interest is the proportion of elderly people with depression. Data will be presented using a forest plot, including individual prevalence, pooled estimates, and 95% confidence intervals (CI). All pooled estimates will be calculated using an appropriate model (fixed or random-effects model meta-analysis), based on the level of heterogeneity. Heterogeneity between studies will be examined using Cochran’s Q test and quantified using the I^2^ statistic. A rough estimate of the heterogeneity will be as follows: I^2^ 0% to 40% - might not be important; I^2^ 30% to 60% - may represent moderate heterogeneity; I^2^ 50% to 90% - may represent substantial heterogeneity; and I^2^ 75% to 100% - considerable heterogeneity. The importance of the observed I^2^ value will depend on (if) magnitude and direction of effects and (ii) strength of evidence for heterogeneity. Sensitivity and subgroup analyses will be used to identify the causes of heterogeneity. If required, meta-regression will be employed to determine the sources of heterogeneity (30).

A cumulative meta-analysis will be done to detect temporal trends in the depression prevalence over the years and the effect of quality of studies. In the cumulative meta-analysis, the studies are added one at a time in a specified order (e.g., according to date of publication), and the results are summarised as each new study is added. In a forest plot of a cumulative meta-analysis, each horizontal line represents the summary of the results as each study is added, rather than the results of a single study (31,32). All analyses will be done using updated versions of STATA (33) and R software (with meta and metafor packages) (34,35).

#### Assessment of publication bias

We will assess the publication bias by visual inspection of funnel plots and testing using Egger’s weighted regression, with p<0.1 considered indicative of statistically significant publication bias (36).

#### Sensitivity analysis

Sensitivity analysis (37) will be done to remove the influence of low-quality studies. We will also explore the effect of small studies (fewer than 100 participants) and the studies not fulfilling age criteria adequately, such as participants aged 65 years or more. In particular, the Leave-One-Out method (also known as Jackknife estimation) in which we recalculate the results of our meta-analysis K−1 times (where K is a total number of studies), each time leaving out one study. We will then compare the new pooled prevalence with that of the original pooled prevalence of depression. If the new pooled prevalence lies outside of the 95% CI of the original pooled prevalence, we will conclude that the excluded study has a significant effect on the pooled estimate and should be excluded from the final analysis (38,39). Some other issues may also be identified for sensitivity analysis during the systematic review process.

#### Analysis of subgroups or subsets

To reduce the random variations between the estimates of primary studies, we will perform subgroup analysis wherever feasible according to study setting, geographical region (states), states by GDP per capita, type of diagnostic instrument, dementia screening, sampling design, and study period.

## Patient and Public Involvement

No patients are directly involved in this study. The data for systematic review will be collected from previously published studies.

## Discussion

Screening tools are simple to administer, take less time, and are highly useful in primary care settings to screen the people for depression (40). However, confirming a diagnosis of depression by a diagnostic tool provides a more accurate picture of the magnitude of depression. Based on earlier literature, the estimated prevalence of depression was significantly higher when self-reporting instruments or screening tools were used to assess depression (25,41). The estimation based on screening tools varied widely with the type of study tools, geographic region, sample size, sampling methods, and prevalent socio-cultural differences in the country. These may be responsible for different levels of mental health disorders in India. Hence, we will address this issue by using different meta-analytic techniques such as subgroup and sensitivity analyses such as jackknife estimation, meta-regression, and cumulative meta-analyses.

In India, the National Mental Health Survey (NMHS) reported a lower prevalence of lifetime depression (3.14%) and during the previous 12-month period (1.7%) (42) compared to pooled data from 18 countries (n= 89,037) which estimated the average lifetime and 12-month prevalence estimates of DSM-IV MDE to be 14.6% and 5.5% in 10 high-income countries, and 11.1% and 5.9% in 8 low-to middle-income countries, respectively (43). This study will provide the unique opportunity to compare the magnitude of depression estimated using screening tools and findings of NHMS with the pooled estimate of various research studies that have used diagnostic instruments for the identification of depression among elderly persons in India.

In India, mental health services receive a minor fraction of the overall health budget, which is grossly inadequate in proportion to the rising burden of mental disorders. Also, there is a lack of robust and reliable data to address the need for community based mental health services planning and management. The findings of this study, i.e., the estimated magnitude of depression among elderly persons using diagnostic instruments, distribution among subgroups, and regions will help to plan and manage geriatric mental health program in a better way and will provide further directions to future research in the depression epidemiology and its burden in the elderly population. It will also strengthen the provision of comprehensive mental health services in primary health care settings, especially, among the geriatric population in India.

## Data Availability

Not applicable

## Ethics and dissemination

Ethical approval is not required as it will be a systematic review. Data from previously published studies will be retrieved and analyzed. Findings will be disseminated through a peer-reviewed publication in a scientific journal and conferences.

## Abbreviations

GDP: Gross Domestic Product

## Supplementary data

Not applicable.

## Availability of data and materials

Will be available once collected.

## Author Contributions

Conceived the idea: PMB, MB, VY, MP, and SDG. Designed the protocol and wrote the paper: MB, VY, MP, PMB, DD, SMB, VM, SDG, and SP. Critical revision to the manuscript: MB, VY, MP, PMB, DD, SP, VM, and SDG. All authors have read and approved the manuscript. PMB and MB are the guarantors of the paper.

## Data sharing statement

No additional data are available.

## Acknowledgments

None.

